# Discovering disease-causing pathogens in resource-scarce Southeast Asia using a global metagenomic pathogen monitoring system

**DOI:** 10.1101/2021.09.25.21262905

**Authors:** Jennifer A. Bohl, Sreyngim Lay, Sophana Chea, Vida Ahyong, Daniel M. Parker, Shannon Gallagher, Jonathan Fintzi, Somnang Man, Aiyana Ponce, Sokunthea Sreng, Dara Kong, Fabiano Oliveira, Katrina Kalantar, Michelle Tan, Liz Fahsbender, Jonathan Sheu, Norma Neff, Angela M. Detweiler, Sokna Ly, Rathanak Sath, Chea Huch, Hok Kry, Rithea Leang, Rekol Huy, Chanthap Lon, Cristina M. Tato, Joseph L. DeRisi, Jessica E. Manning

## Abstract

Understanding the regional pathogen landscape and surveillance of emerging pathogens is key to mitigating epidemics. Challenges lie in resource-scarce settings, where outbreaks are likely to emerge, but where laboratory diagnostics and bioinformatics capacity are limited. Using unbiased metagenomic next generation sequencing (mNGS), we identified a variety of vector-borne, zoonotic and emerging pathogens responsible for undifferentiated fevers in a peri-urban population in Cambodia. From March 2019 to October 2020, we enrolled 473 febrile patients aged 6 months to 65 years of age presenting to a large peri-urban hospital in Cambodia. We collected sera and prepared sequencing libraries from extracted pathogen RNA for unbiased metagenomic sequencing and subsequent bioinformatic analysis on the global cloud-based platform, IDseq. We employed multivariate Bayesian models to evaluate specific pathogen risk causing undifferentiated febrile illness. mNGS identified vector-borne pathogens as the largest clinical category with dengue virus (124/489) as the most abundant pathogen.

Underappreciated zoonotic pathogens such as *Plasmodium knowlesi*, leptospirosis, and co-infecting HIV were also detected. Early detection of chikungunya virus presaged a larger national outbreak of more than 6,000 cases. Pathogen-agnostic mNGS investigation of febrile persons in resource-scarce Southeast Asia is feasible and revealing of a diverse pathogen landscape. Coordinated and ongoing unbiased mNGS pathogen surveillance can better identify the breadth of endemic, zoonotic or emerging pathogens and deployment of rapid public health response.

**Clinical Trial Numbers:** NCT04034264 and NCT03534245.

**Significance Statement:** Public health authorities recently advocated for global expansion of sequencing capacity worldwide; however, the importance of genomics-based surveillance to detect emerging pathogens or variants in resource-limited settings is paramount, especially in a populous, biodiverse Southeast Asia. From 2019 to 2020, pathogen metagenomic next generation sequencing (mNGS) of febrile patients in Cambodia identified several vector-borne and zoonotic pathogens, both common and underappreciated, and resulted in a variety of actionable health interventions. Understanding these pathogen discoveries, and the attendant challenges of mNGS in these outbreak-prone settings, is critical for today’s global society and decision-makers in order to implement sequencing-based pathogen or variant detection.

**Significance Statement:** Metagenomic pathogen sequencing offers an unbiased approach to characterizing febrile illness. In resource-scarce settings with high biodiversity, it is critical to identify disease-causing pathogens in order to understand burden and to prioritize efforts for control. Here, mNGS characterization of the pathogen landscape in Cambodia revealed diverse vector-borne and zoonotic pathogens irrespective of age and gender as risk factors. Identification of key pathogens led to changes in national program surveillance. This study provides a recent ‘real world’ example for the use of mNGS surveillance in both identifying diverse microbial landscapes and detecting outbreaks of vector-borne, zoonotic, and other emerging pathogens in resource-scarce settings.

**Classification:** Biological Sciences; microbiology; medical sciences

## Introduction

A global pathogen surveillance network can best identify emerging and underlying pathogens if it employs pathogen-agnostic detection methods, such as metagenomic next-generation sequencing (mNGS), and is decentralized to include low-resource settings that are often biodiversity hotspots at increased risk for disease outbreaks (1–3). Lack of diagnostics in these areas makes undifferentiated febrile illnesses difficult to diagnose and treat, much less confirm and report for global public health awareness. In Southeast Asia where a quarter of the world’s population resides, rapid but heterogeneous economic development juxtaposes low-resource and high-resource areas, causing high cross-border mobility of persons for economic opportunities. In Cambodia and Laos, laboratory testing for non-malarial fevers is limited, particularly in rural and peri-urban areas where simple diagnostics like dengue rapid tests may not be available (4). In many instances, healthcare providers make diagnoses and empiric treatment decisions based on symptoms so the responsible pathogen is rarely identified.

Syndromic diagnosis is an epidemiological pitfall in Southeast Asia because the true scope of pathogen diversity remains poorly defined. From limited decade-old surveillance data of febrile Cambodians, *Plasmodium* infections made up more than 50% of the responsible pathogens followed by pathogenic *Leptospira* (9.4%), influenza virus (8.9%), and dengue virus (DENV)(6.3%) (5). In a separate serosurvey, one third of febrile Cambodian patients had antibodies to rickettsiae that cause scrub typhus (via chiggers containing *Orientia tsutsugamus*hi), endemic typhus (via rat fleas *Xenopsylla cheopia* carrying *Rickettsia typhi*), spotted fever (via ticks carrying *Rickettsia rickettsii*), and murine typhus (via cat fleas *Ctenocephalides felis* carrying *Rickettsia felis)*, which some speculate may be the next mosquito-borne outbreak (6, 7). Entomological studies of field-collected ticks, mosquitos and fleas in Cambodia have revealed high biodiversity of potential disease-carrying vectors including underappreciated *Bartonella spp* (8, 9). Other serosurveys of bats, domestic pigs, and birds in Cambodia demonstrated the presence of antibodies to other zoonotic viruses including Nipah virus, hepatitis E, Japanese encephalitis virus, and West Nile virus with potential for spillover into the human population (10–12).

In these settings of high pathogen diversity, monitoring with pathogen-agnostic tools, such as mNGS, is ideal but typically not available in-country to provide results within an actionable timeframe. Examples of mNGS identifying pathogens in patients are limited to clinical research programs in developed countries (13–15). However, it is clear that broadly applied and timely mNGS in any population can lead to a better understanding of the overall pathogen landscape, which has direct implications for disease containment methods in the event of an outbreak (16, 17). Here, as an initial step in a low-resource setting in Asia, we describe implementation of mNGS serosurveillance, using an open-source cloud-based bioinformatics tool, to identify pathogens in sera from febrile individuals in peri-urban Cambodia.

## Results

### Clinical characteristics of febrile participants in Cambodia

From March 2019 to October 2020, a total of 487 patients presenting with fever were screened, enrolled and contributed sera for mNGS (377 patients in hospital-based cohort and 110 in community-based cohort) (Figure 1). Demographic and clinical characteristics are detailed in Table 1; notably, the participants are young with the median age in the hospital cohort at 10 years (IQR 12), and 6 years (IQR 4) in the community cohort. The predominant symptom reported in both studies was headache 52.4% (256/487). Of the adults, 67.7% (61/90) were employed in non-agricultural settings while the remainder were farmers or unemployed. In only the hospital cohort, approximately half of participants reported insect exposure, primarily mosquitos (211/376). Nearly three-quarters of participants reported animal exposure (275/376). The most common animal exposures included dogs, cats, and chickens, with some rare reports of exposure to pigs and horses.

**Figure 1.**
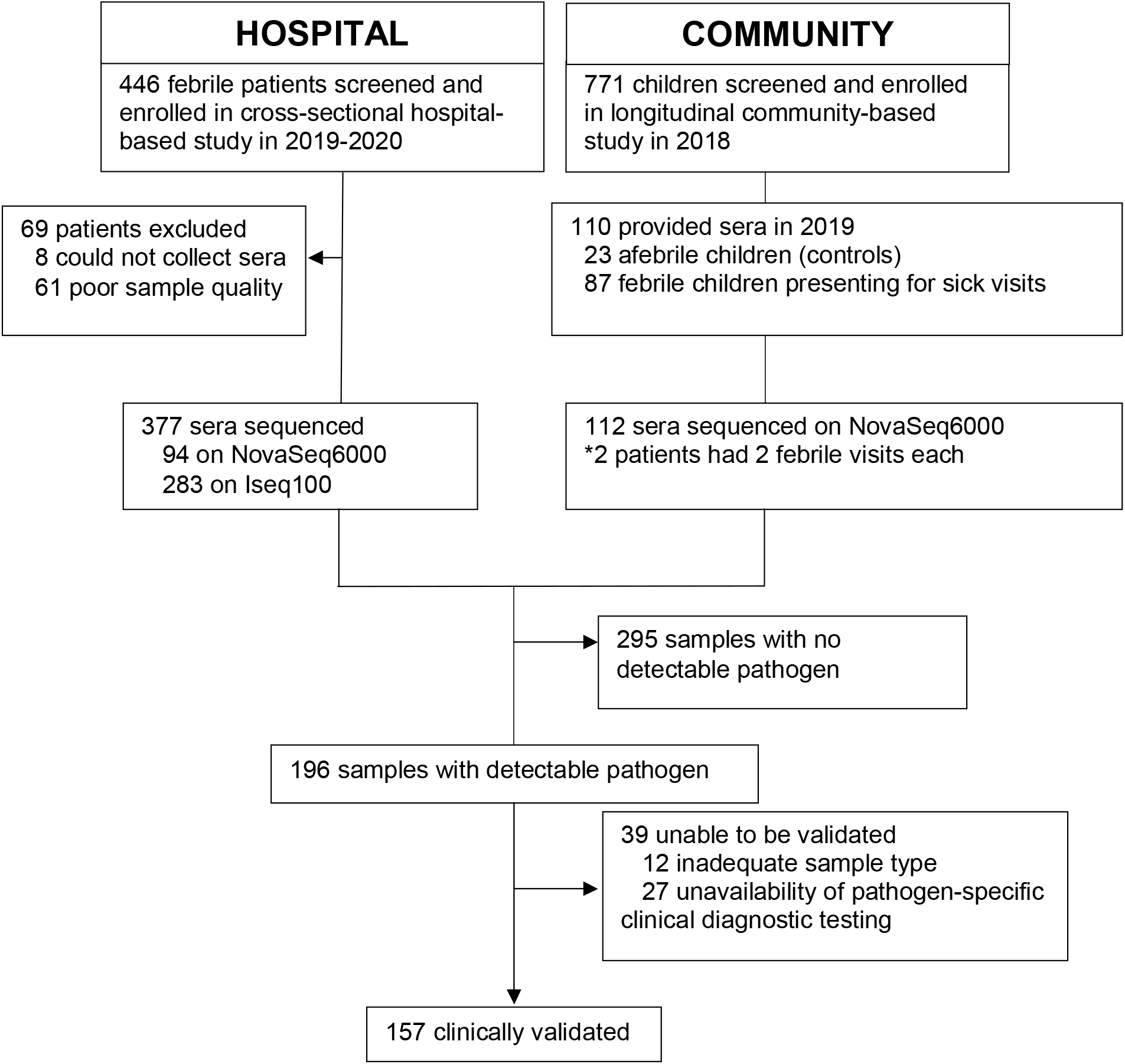
Study flow chart. Flow of enrolled febrile patients through two clinical studies defined as hospital (cross-sectional febrile patient hospital-based cohort) and community (longitudinal pediatric community-based cohort).

**Table 1.**
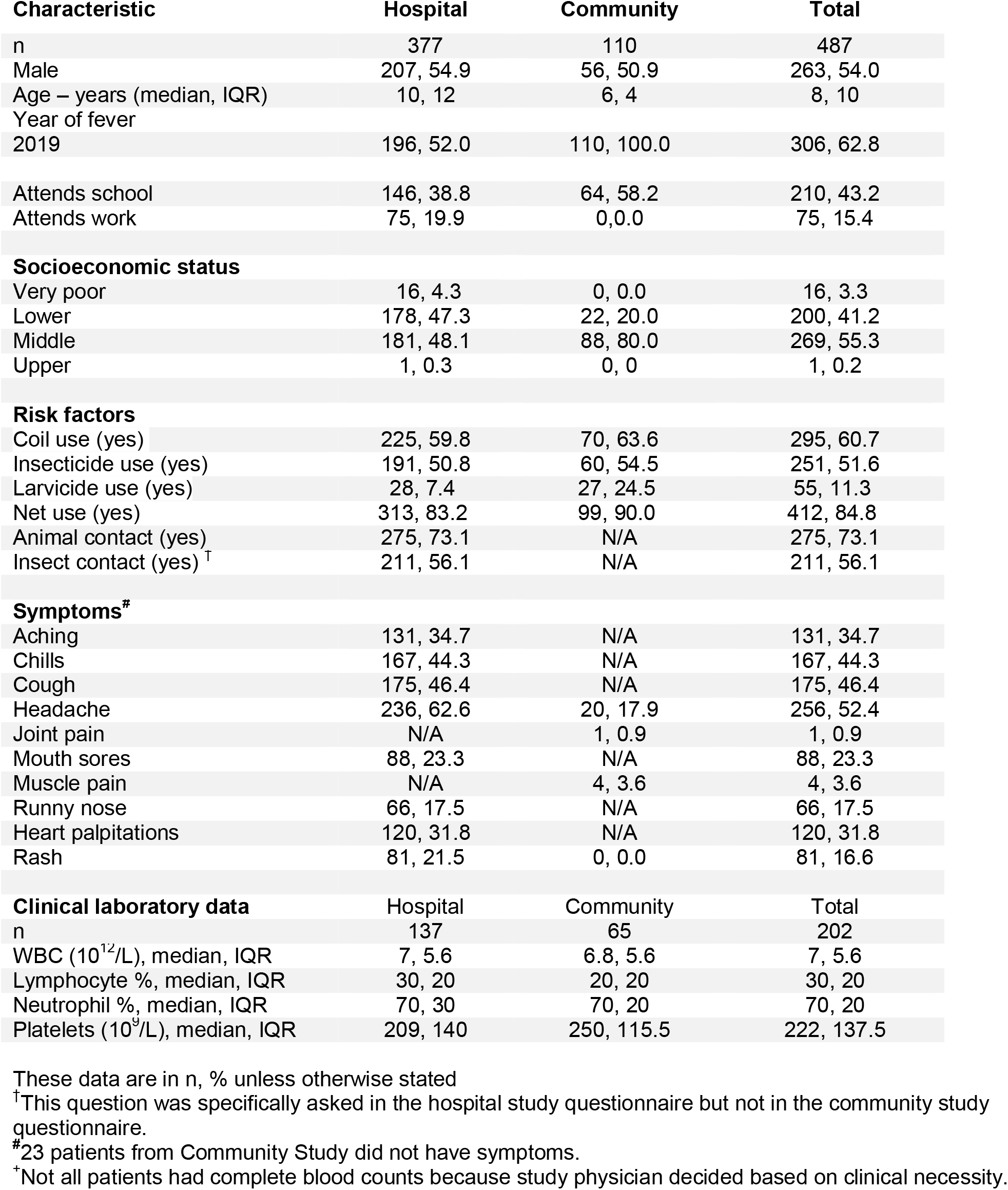
Baseline demographic and clinical characteristics of cohort.

### mNGS Characterization of the Pathogen Landscape in Febrile Cambodians

The composite of identified pathogens in both cohorts is shown in Figure 2. Vector-borne disease was the most prevalent clinical category for mNGS analysis of sera from febrile patients. This clinical category included DENV (138/489), most abundant, followed by rickettsiae (13/489), CHIKV (10/489) and *P. vivax* (6/489). The second highest clinical category was systemic viral illness notably including hepatitis and pegiviruses (8/489).

**Figure 2.**
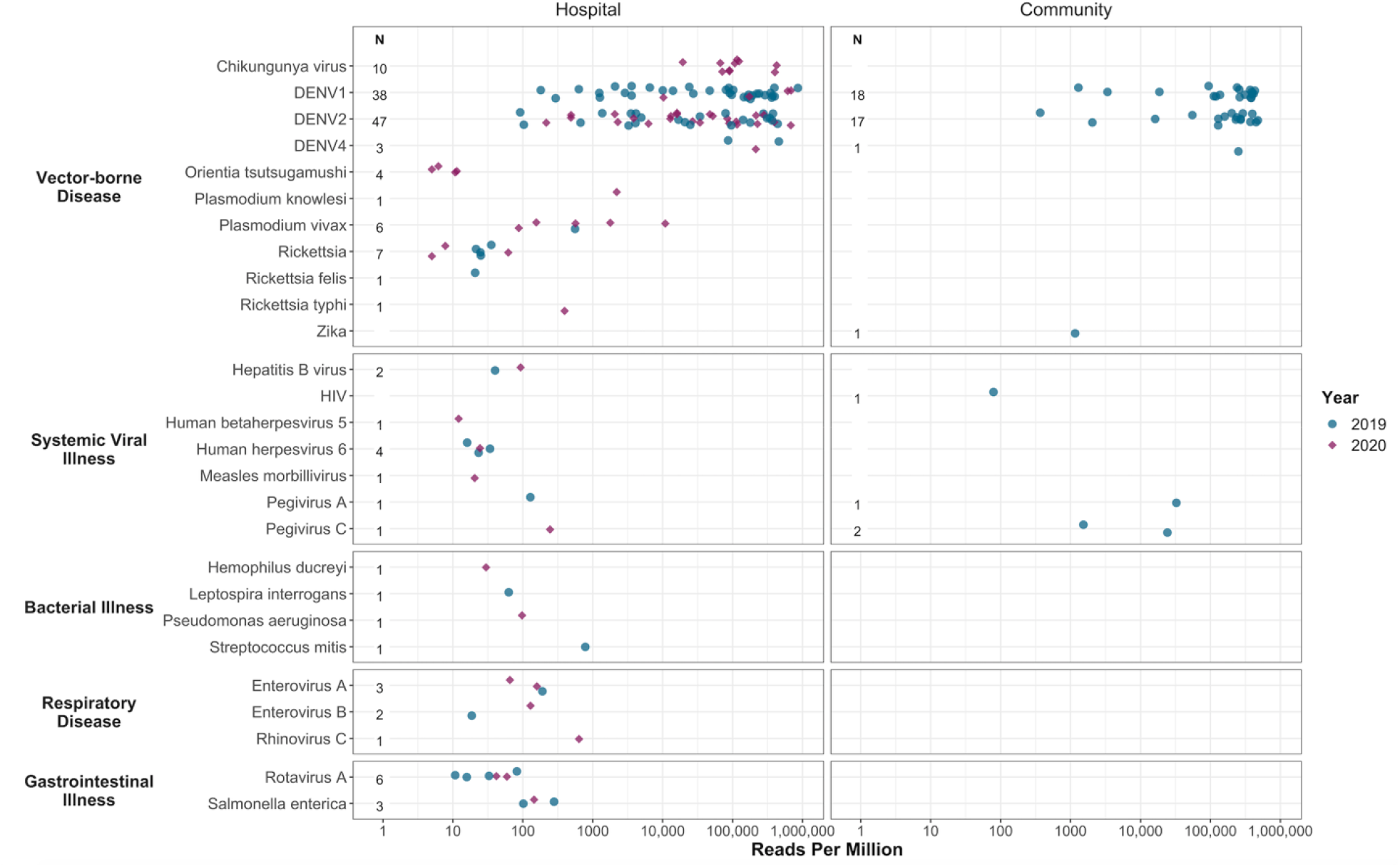
Microbial landscape identified from serum samples of febrile Cambodian participants. Identified pathogens in sera by clinical category, reads per million, and study setting. Each circle represents a pathogen in 2019 and each diamond a pathogen in 2020.

### Pathogen Serosurveillance Findings in Clinical and Regional Contexts

Here, we describe select pathogens in greater detail pertinent to clinical and genomic epidemiology of the Southeast Asian region.

#### Dengue virus

Dengue was responsible for the greatest disease burden in our study (138 DENV positive cases of 489 febrile cases) due to the largest DENV outbreak documented in Cambodian history in 2019. The predominant DENV serotype of the outbreak in Kampong Speu province was DENV1 (Supplemental Table 2). 71% (48/67) of DENV1 sequences identified, aligned to DENV1, accession no. MF033254.1 from 2016 DENV1 outbreak in Singapore. Phylodynamic analyses will be presented elsewhere.

#### Rickettsia

While rickettsial diseases are easily treated with oral doxycycline, the challenge is timely diagnosis and access to serological and/or molecular testing for confirmation. In Laos, a country of similar climate and socio-economic status as Cambodia, 7% (122/1871) of febrile patients were positive for scrub typhus, 1% murine typhus (10/1849) and 1% undetermined *Rickettsia spp*. combined with *Rickettsia felis* (9/1849) (21). Here, four patients were positive for *Orientia tsutsugamushi*, highly homologous to accession no. CP044031.1 from Zhejiang province, China and to accession no. LS398552.1 from Udon Thani, Thailand (22). mNGS identified one case of *Rickettsia felis*, one of *Rickettsia typhi* and seven cases of the genus *Rickettsia* without clinical confirmation of species-level data.

#### Chikungunya virus

In July 2020, we identified 10 cases of CHIKV in Kampong Speu Province where patients presented with symptoms of fever, rash, shaking chills and arthralgias. mNGS analysis revealed CHIKV as the clinical etiology after initial diagnoses of DENV were made based on patients’ presenting symptoms. These sequences aligned closely with three Urban Asian Lineage (AUL) sequences from Thailand (accession nos. MN075149.1, MN630017.1 and MK468801.1). CHIKV PCR was then added to national surveillance and it was noted that the outbreak spread rapidly to 21 other provinces in Cambodia, affecting at least 6,000 people by the end of September 2020 despite implementation of vector control (23).

#### Zika virus

ZIKV circulates at low levels in Thailand and Vietnam, however almost no active cases have been reported in Laos and Cambodia even during the global epidemic in 2015-16.(24, 25) Since 2010, only one prospective case of active ZIKV infection was detected in Cambodia, notably Kampong Speu province.(26) In the current study, sera from an otherwise asymptomatic school-aged female was positive for ZIKV with 20.1x coverage depth and 98.8% coverage breadth closely aligned with to accession no. MF996804.1, a Thai case of microcephaly, with 99.2% sequence similarity. These information indicate that ZIKV in Cambodia has regional sequence similarities to Thailand, possibly related to high cross-border traffic between the two countries despite little ZIKV detected in Cambodia.(27) Another possibility is a separate enzootic ZIKV transmission cycle maintained in non-human primates given recent evidence of ZIKV in stump-tailed macaques in Thailand (28).

#### Plasmodium spp

Cambodia is in the pre-elimination stage for all malarial species with a specific goal to eliminate *P. falciparum* by 2025 (29). mNGS identified six cases of very low parasitemia (down to 16 parasites per uL) with *P. vivax*, initially missed on microscopy or rapid test. *P. vivax* has replaced *P. falciparum* as the most prevalent form of malaria in Southeast Asia, particularly in Cambodia where eradicative liver-stage treatment of *P. vivax* with primaquine has not yet been widely adopted (4). mNGS also identified *P. knowlesi* in a forest worker, previously diagnosed with *P. malariae* using blood smear microscopy. This pathogen identification led to retrospective mNGS assessment of other *P. malariae* cases and the addition of *P. knowlesi* PCR to national surveillance. Given human encroachment and deforestation in Southeast Asia, there is ample opportunity for spread of zoonotic malaria, such as *P. knowlesi* typically found in non-human primates, that may endanger elimination goals (4).

#### Leptospira interrogans

Leptospirosis is an underappreciated health threat in Southeast Asia. In nearby Kampong Cham province, 2.5% (17/630) of all fevers in 27 rural to semi-rural villages were confirmed as acute leptospirosis infection via IgM serology and microagglutination testing (30). In November 2019, a school-aged female with a fever of 38.5°C presented with a headache and abdominal pain. mNGS identified *Leptospira interrogans* at 62.9 rpM with 99.7% homology to CP048830.1. Due to limited in-country diagnostic testing, no further testing was performed, but clinical examination confirmed the presence of conjunctival effusion, a specific feature of leptospirosis.

#### HIV-1 and DENV co-infection

A school-aged female of Vietnamese descent presented to the hospital with a 39°C fever and mNGS analysis revealed a possible coinfection of DENV2 and HIV. The low sequence coverage (14%) of a Vietnamese HIV genome, accession no. FJ185253.1, was likely due to the sequencing space used on the high number of DENV2 reads (DENV2: 368 rpM, 99% sequence coverage breadth and 17.1x depth versus HIV: 78.1 rpM, 14% coverage and 1x depth) for this sample (31). However, re-mapping all reads belonging to the *Lentivirus* genus resulted in a more comprehensive assessment with 33% coverage of the Vietnamese HIV-1 viral genome (accession no. FJ185246. 1) at a depth of 3.47x, with greatest homology to a Thai HIV-1 strain, accession no. LC114832.1, from a female sex worker. The mNGS results were confirmed by clinically validated HIV 1/2 antibody tests, and the patient subsequently initiated antiretroviral therapy.

### Risk modeling of contracting vector-borne disease

The probability of having a vector-borne infection was increased for individuals in the hospital study if the household owned a car, (OR 1.95, 95% 1.19–3.21) or if they were 5 to 18 years of age (for 5 to 10 years of age; OR 2.35, 95% CI (1.11–5.06); OR 2.68 for 10 to 18 years of age; 1.4–5.29) (Figure 3; Table 2). In the community cohort, living near surface flooding, using a Scaled Flooding Index (4-week lag), significantly increased the likelihood of vector-borne infection (OR 2.04, 95% CI 1.24–3.49) while larvicide use decreased the chances of acquiring a vector-borne disease (OR 0.32 95% CI 0.11–0.8).

**Figure 3.**
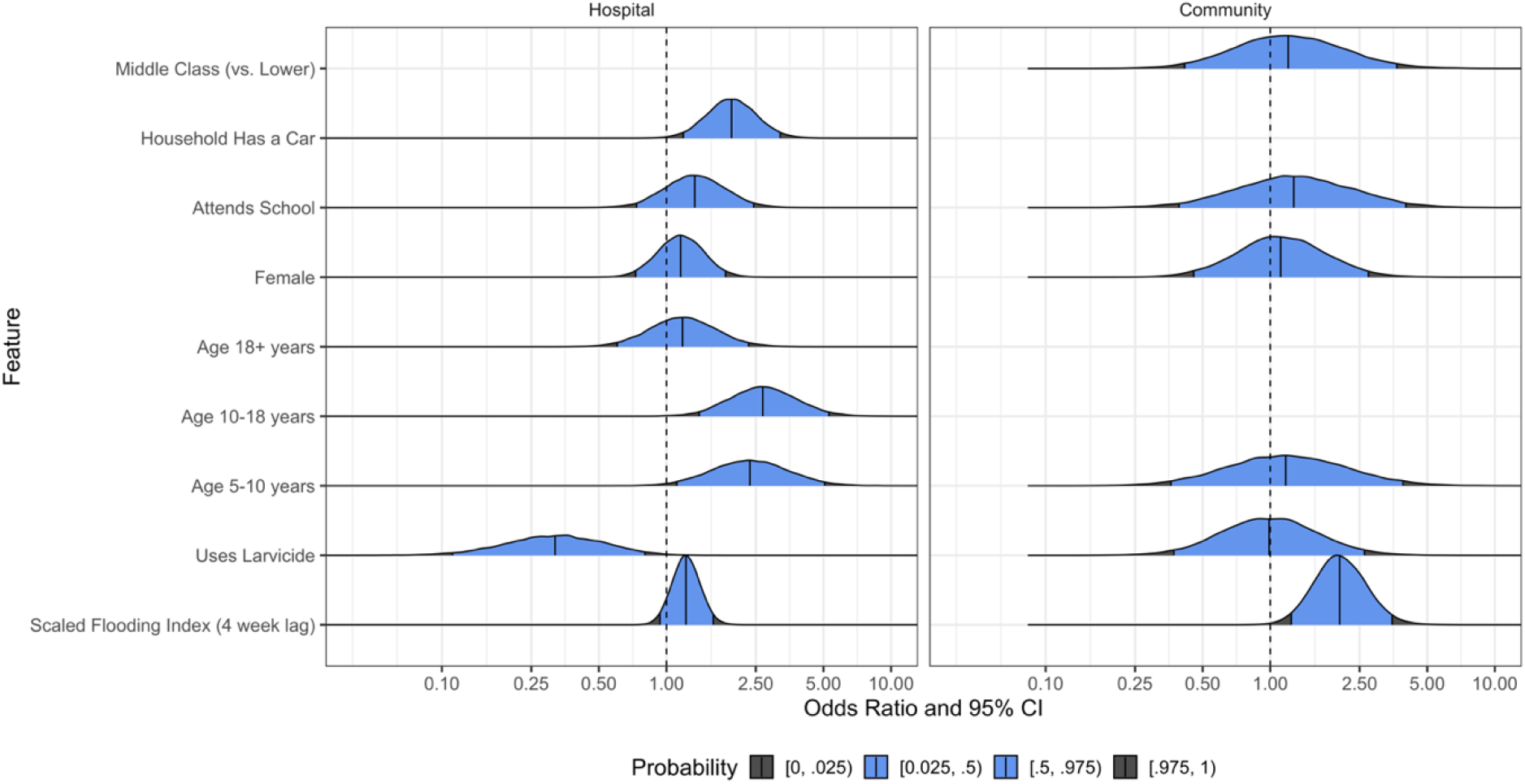
Probability of infection by vector-borne pathogen. Results of multivariate analyses in both patient populations to identify risk factors of contracting vector-borne pathogens.

**Table 2.** Predictors for infection by vector borne versus non-vector-borne pathogens

| Risk Factors |  | Hazard ratio | 95% CI |
| --- | --- | --- | --- |
| Scaled flooding index |  |  |  |
|  | Hospital | 1.22 | 0.94 - 1.61 |
|  | Community | 2.04 | 1.24-3.49 |
| Uses larvicide |  |  |  |
|  | Hospital | 0.32 | 0.11-0.8 |
|  | Community | 0.99 | 0.37-2.63 |
| Age 5–10 years |  |  |  |
|  | Hospital | 2.35 | 1.11-5.06 |
|  | Community | 1.17 | 0.36-3.9 |
| Age 10–18 years |  |  |  |
|  | Hospital | 2.68 | 1.4-5.29 |
|  | Community | N/A | N/A |
| Age 18+ years |  |  |  |
|  | Hospital | 1.18 | 0.6-2.32 |
|  | Community | N/A | N/A |
| Female |  |  |  |
|  | Hospital | 1.16 | 0.73-1.83 |
|  | Community | 1.11 | 0.46-2.74 |
| Attends school |  |  |  |
|  | Hospital | 1.34 | 0.74-2.44 |
|  | Community | 1.27 | 0.39-4.02 |
| Household has a car |  |  |  |
|  | Hospital | 1.95 | 1.19-3.21 |
|  | Community | N/A | N/A |
| Middle class (vs. lower) |  |  |  |
|  | Hospital | N/A | N/A |
|  | Community | 1.2 | 0.42-3.66 |

Crop land was the predominant land cover type for participants’ homes (89%; 426/476; Figure 4) with urban as the next most common (10%; 49/476). Urban participants were more likely to have non-vector borne diseases (13%; 4/30) than vector-borne pathogens (9%; 15/162); however, there were still participants from primarily urban areas with CHIK, DENV1, DENV2, or ZIKV infections (Supplemental Table 1). Interestingly, 92% (125/135) of DENV cases were from crop land (Supplemental Table 2). Formal analyses by each disease outcome were not pursued because of small overall counts. Exploratory univariate analysis of the EIs indicated that only the surface flooding index was associated with any of the disease outcomes (Figure S1).

**Figure 4.**
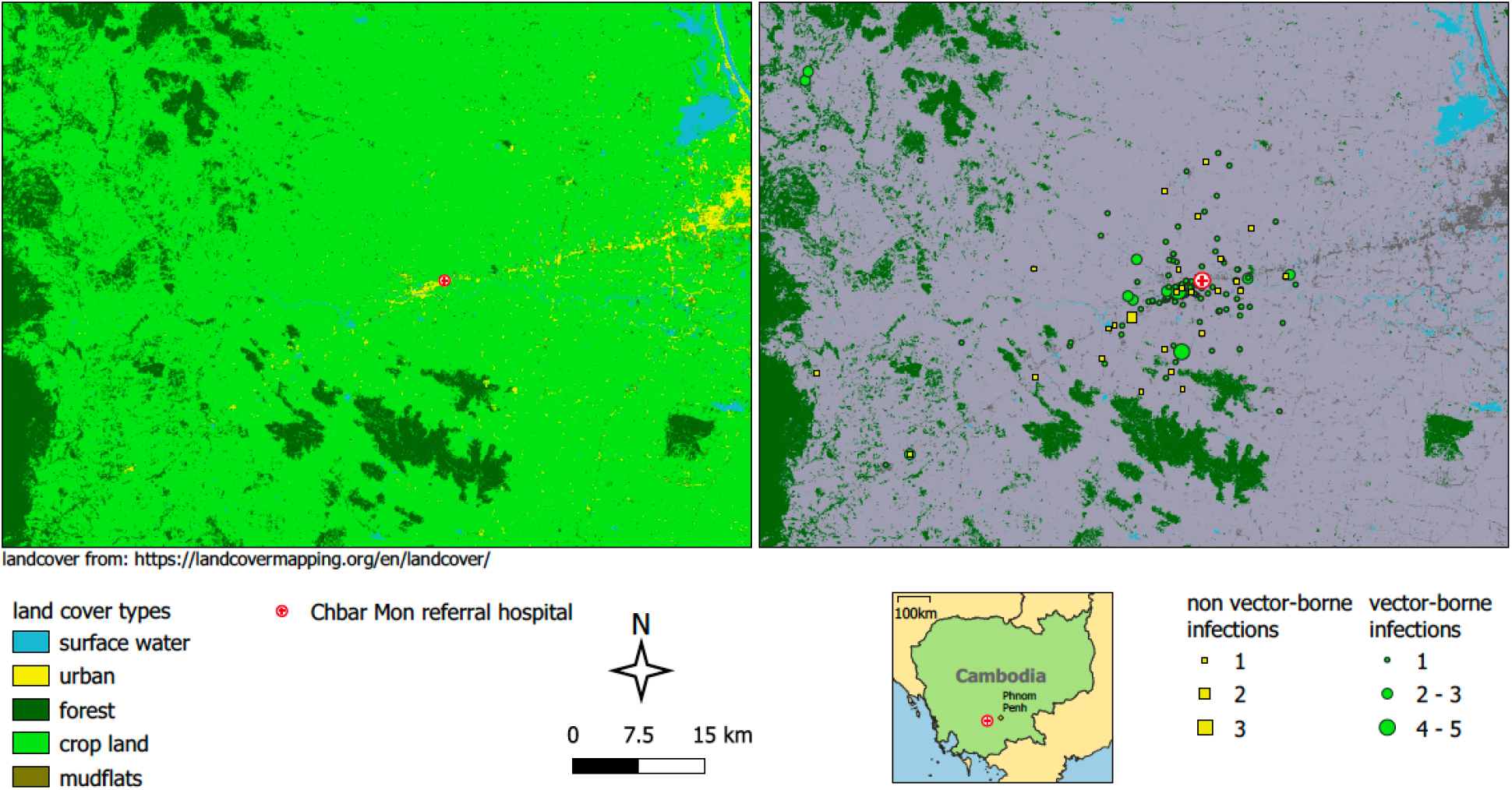
Study site and land use map. Patient locations classified by land use and vector-borne disease status.

## Discussion

Metagenomic NGS serosurveillance from peri-urban Cambodia revealed a diverse pathogen landscape, rich in underappreciated vector-borne and zoonotic pathogens, responsible for febrile disease. In this prospective, cross-sectional mNGS study, we identified common and confounding pathogens and demonstrated the feasibility and usefulness of a decentralized metagenomic sequencing pipeline. Despite challenges of actionable mNGS surveillance in a resource-scarce settings, we contributed to genomics-informed pathogen epidemiology that is otherwise lacking in Cambodia and other similar settings, yet globally relevant given major demographic and socioeconomic shifts underway in the region that may increase the likelihood for disease epidemics (4, 32).

Our study provides a more granular analysis of changing pathogen dynamics than prior surveillance with pre-determined targeted diagnostics like PCR (5,33,34). The hierarchy of species abundance identified here is likely attributed to current malaria elimination campaigns, heterogenous socioeconomic development, and increased human migration (4, 35). DENV is now the most prevalent pathogen, particularly in children in comparison to neighboring countries, while malaria makes up a less substantial portion of febrile cases than in prior years where PCR-confirmed malaria infections was as high as 51.1% (754/1475) of febrile individuals presenting to peri-urban hospitals (5, 32).

Over the past two decades, the importance of vector-borne pathogens as drivers of epidemics and as emerging pathogens cannot be discounted despite current threats posed by novel respiratory pathogens. The detection of primarily vector-borne pathogens in this study is relevant as genomic surveillance becomes the foundation of global health security. The Asian strain of ZIKV evolved to enhance infectivity of humans and mosquitos via a single alanine-to-valine substitution that increased NS1 antigenemia, ultimately resulting in epidemics as early as 2007 in Micronesia and later in the Americas linked to microcephaly (36). Prior CHIK outbreaks were traced to a single mutation in 2005 allowing increased fitness of CHIK in *Aedes albopictus* mosquitos, and thus conferring epidemic potential of the virus in humans (37). Today, autochthonous CHIK transmission and outbreaks occur in increasingly warmer temperate zones like Europe (38). These two separate arboviral mutations, each responsible for devastating epidemics of global impact, highlight the importance of expanding genomic epidemiology and surveillance of vector-borne pathogens. Furthermore, mNGS recently identified novel vector-borne pathogens including the tick-borne flaviviruses like Alongshan and highly fatal mosquito-borne orthobunyaviruses like Cristoli, Umbre, and others (39–41). These emerging pathogens were identified in high-resource areas where clinical staff had access to mNGS technology. This further highlights the importance of real-time, in-country metagenomic investigation of potential pathogens of concern. Logistical and bureaucratic delays in shipping samples out of a country may translate to the establishment and spread of a pathogen in the interim.

To that end, timely contribution of pathogen genomic information from resource-limited settings is critical to the future success of pathogen identification based on genomic sequence data in an increasingly connected world, exemplified by GISAID and GENBANK during the SARS-CoV-2 pandemic (3). The lack of publicly available sequence data of clinically relevant pathogens, such as DENV and CHIK in Southeast Asia, is stark given the regional magnitude of infections by these pathogens.

Our mNGS surveillance primarily identified vector-borne pathogens; therefore, our risk models aimed to inform deductive algorithms for undifferentiated fevers in the region. Judicious use of mNGS surveillance would not entail sequencing every undifferentiated fever that is presumed to be dengue. With dengue being the most common diagnosis attributed to fevers in pediatric patients, we aimed to include demographic, behavior, and ecological data that might stratify risk of a vector-borne disease pathogen versus other pathogens in the hospital-based cohort of all ages. Exposure to animals and occupations did not stratify to any risk, but younger age, household car ownership (a surrogate of socioeconomic status) and absence of larvicide use led to increased risk of vector-borne diseases. Advances in land cover analysis now permit disease risk assessment of a population based on their environment. Here, living near surface flooding increased vector-borne disease risk, and surprisingly, DENV cases originated primarily in crop zones that often border urban zones, corroborating previous claims that DENV transmission in Southeast Asia is both rural and urban (42). Even with these tools to aid diagnostic algorithms, it is evident from our data that assigning microbial etiology to undifferentiated fever based on symptoms and demographic data is difficult given the presence of diverse pathogens, the shifting of socioeconomic patterns, and the ongoing transformation of land cover.

Limitations in the study included the sampling strategy of sera or whole blood alone, primarily for operational purposes in the early establishment of this pathogen mNGS detection pipeline. To that point, exclusive use of sera contributed to our pathogen detection rate of 40%, likely overestimating vector-borne pathogens to the detriment of respiratory and gastrointestinal pathogens. Since completion of the data analysis presented here, our mNGS monitoring efforts now include nasopharyngeal swabs in addition to ongoing blood sampling. To date, the addition of nasopharyngeal sampling to our mNGS surveillance study has led to timely recovery of entire SARS-CoV-2 genomes, with and without enrichment, for variant identification (43, 44). Fortunately, genome recovery of most viruses was straightforward from sera, but sampling limitations remain for other taxa; for example, the optimal sample type to identify and speciate *Rickettsiae* is buffy coat, as opposed to sera, because the bacteria are intracellular (45). Other challenges included identification of less abundant bacterial pathogens, attributable to limited coverage offered by the iSeq, variable host contamination, different library preparation (e.g. DNA-based instead of RNA-based), and again, sample type. The cross-sectional study design limited our ability to see if a patient’s clinical course evolved over a longer period of time, and the lack of blood culture capabilities at this hospital did not allow comparison of mNGS to standard diagnostic techniques for bacterial pathogen identification. However, we strived for actionable data, from either a clinical or public health standpoint, and succeeded in cases of *Plasmodium* spp., HIV, CHIK, and other pathogens. The cost of sequencing is declining while the efficiency of sequencing workflows is increasing, but mNGS analysis of pathogens is still more expensive than targeted diagnostics like PCR or culture (1). Until now, the majority of sequencing and analysis of biological samples collected in Cambodia and other resource-limited settings was outsourced to the Global North. To overcome challenges in reagent procurement, internet connectivity, and lack of advanced bioinformatics training, we built a robust infrastructure to mitigate these issues while also relying upon a pre-curated, rapid bioinformatics pipeline to build in-country expertise that allowed the entire sample collection, processing, and mNGS analysis to happen in a public Cambodian laboratory.

As a result, our ongoing, in-country metagenomic sequencing pipeline and capacity-building provides continuous monitoring of common and emerging pathogens for actionable interventions when possible. While the world looks to bolster real-time, genomics-informed pathogen surveillance networks to monitor COVID-19 variants and other emerging pathogens, challenges remain to establish critical “nodes” in biodiverse, resource-scarce areas (3, 46). Yet, as shown here, mNGS pathogen surveillance in these settings is feasible, revealing of diverse microbial landscapes, and paramount to the future of global health security.

## MATERIALS AND METHODS

### Enrollment

Participants living in Kampong Speu province, Cambodia, were eligible for enrollment in: 1) a longitudinal, community-based cohort of children, two to nine years of age (referred to as “community” and considered semi-active surveillance because study participants were told to notify study coordinator when they have a fever, called a “sick visit” that was considered nested cross-sectional timepoint within the longitudinal cohort); and 2) a cross-sectional hospital-based febrile cohort established in July 2019 (referred to as “hospital” and considered passive surveillance because patients first presented to the hospital with fever and were then asked to participate). Overall, participants were required to 1) be 6 months to 65 years of age; and 2) have a measured fever equal to or greater than 38°C in previous 24 hours (see clinicaltrials.gov for full criteria). Demographics, clinical, and risk factor data was stored in a REDCAP® database. Locational data was collected using Garmin® GPS devices and Google Earth.

### Sample collection and Nucleic Acid Extraction

At enrollment, approximately 5 ml of whole blood was collected (except 2ml collected from those under 2 years old). Sera was isolated and stored in cryovials with an equal volume of 2x DNA/RNA Shield (Zymo Research, Irvine, CA) at -20°C and transported from the Kampong Speu Hospital laboratory to the Cambodian National Center for Parasitology Entomology and Malaria Control (CNM) in Phnom Penh, Cambodia. Pathogen RNA was isolated from sera using Quick-RNA MicroPrep Kit (Zymo Research, Irvine, CA) and DNAse-treated.

### Library Preparation

mNGS libraries were prepared from isolated pathogen RNA and converted to cDNA Illumina libraries using the NEBNext Ultra II RNA Library Prep Kit (New England BioLabs, Ipswich, MA). Human rRNA was depleted via FastSelect -rRNA HMR (Qiagen, Germantown, MD). ERCC Spike-In Controls (ThermoFisher, Waltham, MA) were used to indicate potential library preparation errors and to calculate input RNA mass. The initial samples (n=208) were sequenced on a NovaSeq6000 (Illumina, San Diego, CA) instrument as part of a pilot wet lab training at the Chan Zuckerberg BioHub in San Francisco, CA, and then the remainder of the study (n=279) was performed on an iSeq100 (Illumina) in Phnom Penh, Cambodia, using 150 nucleotide paired-end sequencing. Water controls were included in each library preparation.

### Bioinformatic analysis

Raw fastq files were uploaded to the IDseq portal, a cloud-based, open-source bioinformatics platform, to identify microbes from metagenomic data (https://idseq.net).(18) Potential pathogens were distinguished from commensal flora and contaminating microbial sequences from the environment by establishing a Z-score metric based on a background distribution derived from 16 non-template control libraries. Data were normalized to unique reads mapped per million input reads for each microbe at both species and genus levels. Taxa with Z-score less than 1, an average base pair alignment of less than 50 base pairs, an e-score less than 1e-10 and reads per million (rpM) less than 10 were removed from analysis.

### Clinical validation

Pathogens for which clinical testing capabilities were available in-country were validated to include RT-PCR of Hepatitis B, *Plasmodium spp*., DENV, CHIK and ZIKV, serology of Human immunodeficiency virus (HIV) 1/2 antibodies or blood smear examination of *Plasmodium* infections by World Health Organization (WHO)-certified microscopists. Validation testing for other pathogens is underway or being developed. Samples were considered to have ‘no pathogen hit’ if they meet QA/QC standards but no resulting pathogenic organisms were identified with appropriate thresholds in place.

### Spatial and environmental data

Land cover data for Cambodia were downloaded from Open Development Cambodia (https://opendevelopmentcambodia.net). The data come from the Regional Land Cover Monitoring System at a resolution of 30 m by 30 m and were from 2016 (the most recent year we could find at this resolution). We used open-source satellite imagery (Google Earth) to ensure that the land cover data matched the reality on the ground. Participant village locations were then plotted on top of the land cover map. To summarize and quantify land cover types, we created 1km buffers around the geographic coordinates for participant villages and extracted land cover characteristics for each participant using the Zonal Histogram function in QGIS (version 3.16.5: https://qgis.org). We then categorized each participant according to the land cover type that predominated around their village location, and tabulated land cover types according to disease outcomes. Environmental indices (EI) for surface water and vegetation were extracted from Moderate Resolution Imaging Spectroradiometer (MODIS) products (MOD13Q1/MYD13Q1 250 meter AQUA/TERRA 16-day composites). A normalized flooding index (NDFI), the normalized differential vegetation index (NDVI), and the enhanced vegetation index (EVI) were all extracted for this analysis.(19)(20) NDFI gives an indication of surface water, NDVI gives an indication of surface vegetation, and EVI is an improvement on NDVI in that it is less sensitive to atmospheric conditions and forest canopies. The data were downloaded for each 16-day time interval (from July 2018–May 2020) using a 1km buffer around the home of each patient in the dataset. The visit date of each participant was then used to align the EI values for each participant. EI values from the 16-day period leading up to a participant visit were used for analyses.

### Statistical analysis

The primary endpoint is identification of pathogen sequences via IDseq analysis in serum samples from febrile individuals treated at the Kampong Speu District Referral Hospital. On average, we found 25-40% of the monthly febrile cases were attributable to vector-borne disease. As such, we decided to determine which demographic variables, risk factors, and climate data were associated with vector-borne pathogen identification using a Bayesian logistic regression model. For our feature coefficients, we used a weakly informative prior and a MCMC sampler to determine the posterior distribution of the coefficients. We plot the marginal coefficient densities and display the posterior medians along with 95% credible intervals. We fit two separate models: one for the hospital cohort and one for the community cohort. Most, but not all, features are present in both models. More details about variable selection, model diagnostics, and model sensitivity may be found in the supplemental material.

### Data Availability

All genome sequence data from this study have been submitted to the NCBI Sequence Read Archive under Bioproject ID PRJNA681566. All protocols are uploaded on protocols.io and all bioinformatics code is available on https://github.com.

### Ethics

The study protocol was approved by the institutional review boards at the US National Institutes of Health and the National Ethics Committee on Human Research in Cambodia (NCT04034264 and NCT03534245 on clinicaltrials.gov). All individuals provided informed consent. The guardians of all pediatric participants provided signed informed consent to participate in the study; and those aged 14 – 17 also provided assent in addition to parental consent.

## Data Availability

The authors confirm that the data supporting the findings of this study are available within the article [and/or] its supplementary materials and

## Acknowledgements

We thank patients and families of Kampong Speu District Referral Hospital who participated in this study. We thank the Provincial Health Department of Kampong Speu province in Cambodia. We thank all the other employees at the Chan Zuckerberg Biohub and Chan Zuckerberg Initiative not listed in the author byline. We thank Brian Moyer and the NIAID Office of Cyberinfrastructure and Computational Biology (OCICB) for their assistance in improving the cyberinfrastructure of our Cambodian field sites.

## Funding

This research is supported by the Division of Intramural Research at the National Institute of Allergy and Infectious Diseases at the National Institutes of Health and the Bill and Melinda Gates Foundation [grant number OPP1211806]. The authors declare no competing interests.

## Notes

### Competing Interest Statement

The authors have declared no competing interest.

### Clinical Trial

NCT04034264 and NCT03534245

### Author Declarations

The study protocol was approved by the institutional review boards at the US National Institutes of Health and the National Ethics Committee on Human Research in Cambodia (NCT04034264 and NCT03534245 on clinicaltrials.gov). All individuals provided informed consent. The guardians of all pediatric participants provided signed informed consent to participate in the study; and those aged 14 to 17 also provided assent in addition to parental consent.

## References

1. G. L. Armstrong, et al., Pathogen Genomics in Public Health. N. Engl. J. Med. 381, 2569–2580 (2019).

2. X. Deng, et al., Metagenomic sequencing with spiked primer enrichment for viral diagnostics and genomic surveillance. Nat. Microbiol., 1–12 (2020).

3. J. L. Gardy, N. J. Loman, Towards a genomics-informed, real-time, global pathogen surveillance system. Nat. Rev. Genet. 19, 9–20 (2018).

4. R. C. Christofferson, et al., Current vector research challenges in the greater Mekong subregion for dengue, Malaria, and Other Vector-Borne Diseases: A report from a multisectoral workshop March 2019. PLoS Negl. Trop. Dis. 14, e0008302 (2020).

5. T. C. Mueller, et al., Acute undifferentiated febrile illness in rural Cambodia: a 3-year prospective observational study. PloS One 9, e95868 (2014).

6. C.M. Farris et al. Rickettsial Disease: Important causes of undifferentiated Fever in Cambodia. 9th Tick Tick-Borne Pathog Conf Asia Pac Rickettsia Conf 2017;Cairns, Australia.

7. P. Parola, D. Musso, D. Raoult, Rickettsia felis: the next mosquito-borne outbreak? Lancet Infect. Dis. 16, 1112–1113 (2016).

8. D. Prasetyo et al. Bartonellosis in Cambodia and Lao People’s Democratic Republic. 9th Tick Tick-Borne Pathog Conf Asia Pac Rickettsia Conf 2017; Cairns, Australia.

9. S. Boyer, S. Marcombe, S. Yean, D. Fontenille, High diversity of mosquito vectors in Cambodian primary schools and consequences for arbovirus transmission. PLOS ONE 15, e0233669 (2020).

10. J.-M. Reynes, et al., Nipah Virus in Lyle’s Flying Foxes, Cambodia. Emerg. Infect. Dis. 11, 1042–1047 (2005).

11. Y. E. Raji, O. P. Toung, N. M. Taib, Z. B. Sekawi, A systematic review of the epidemiology of Hepatitis E virus infection in South – Eastern Asia. Virulence 12, 114–129 (2021).

12. H. Auerswald, et al., Serological Evidence for Japanese Encephalitis and West Nile Virus Infections in Domestic Birds in Cambodia. Front. Vet. Sci. 7 (2020).

13. M. R. Wilson, et al., Clinical Metagenomic Sequencing for Diagnosis of Meningitis and Encephalitis. N. Engl. J. Med. 380, 2327–2340 (2019).

14. W. Gu, S. Miller, C. Y. Chiu, Clinical Metagenomic Next-Generation Sequencing for Pathogen Detection. Annu. Rev. Pathol. 14, 319–338 (2019).

15. T. Doan, et al., Illuminating uveitis: metagenomic deep sequencing identifies common and rare pathogens. Genome Med. 8, 90 (2016).

16. A. Ramesh, et al., Metagenomic next-generation sequencing of samples from pediatric febrile illness in Tororo, Uganda. PLOS ONE 14, e0218318 (2019).

17. S. Saha, et al., Unbiased Metagenomic Sequencing for Pediatric Meningitis in Bangladesh Reveals Neuroinvasive Chikungunya Virus Outbreak and Other Unrealized Pathogens. mBio 10 (2019).

18. K. L. Kalantar, et al., IDseq-An open source cloud-based pipeline and analysis service for metagenomic pathogen detection and monitoring. GigaScience 9 (2020).

19. Rouse J, Hass R, Monitoring the vernal advancement and retrogradation (Green wave effect) of natural vegetation. NASA-CR-139243 Report No.: E74-10676, 8–9 (1974).

20. M. Boschetti, F. Nutini, G. Manfron, P. A. Brivio, A. Nelson, Comparative Analysis of Normalised Difference Spectral Indices Derived from MODIS for Detecting Surface Water in Flooded Rice Cropping Systems. PLOS ONE 9, e88741 (2014).

21. M. Mayxay, et al., Causes of non-malarial fever in Laos: a prospective study. Lancet Glob. Health 1, e46–e54 (2013).

22. S. D. Blacksell, et al., Genetic typing of the 56-kDa type-specific antigen gene of contemporary Orientia tsutsugamushi isolates causing human scrub typhus at two sites in north-eastern and western Thailand. FEMS Immunol. Med. Microbiol. 52, 335–342 (2008).

23. V. O. D. English, Chikungunya Spreads to 21 Provinces, Almost 6,000 Suspected Infected. Cambodia Dly. (2020) (March 16, 2021).

24. K. Ruchusatsawat, et al., Long-term circulation of Zika virus in Thailand: an observational study. Lancet Infect. Dis. 19, 439–446 (2019).

25. V. Duong, et al., Low Circulation of Zika Virus, Cambodia, 2007–2016. Emerg. Infect. Dis. 23, 296–299 (2017).

26. V. Heang, et al., Zika Virus Infection, Cambodia, 2010. Emerg. Infect. Dis. 18, 349–351 (2012).

27. T. Wongsurawat, et al., Case of Microcephaly after Congenital Infection with Asian Lineage Zika Virus, Thailand. Emerg. Infect. Dis. 24 (2018).

28. D. Tongthainan, et al., Seroprevalence of Dengue, Zika, and Chikungunya Viruses in Wild Monkeys in Thailand. Am. J. Trop. Med. Hyg. 103, 1228–1233 (2020).

29. S. Siv, et al., Plasmodium vivax Malaria in Cambodia. Am. J. Trop. Med. Hyg. 95, 97–107 (2016).

30. S. Hem, et al., Estimating the Burden of Leptospirosis among Febrile Subjects Aged below 20 Years in Kampong Cham Communities, Cambodia, 2007-2009. PLoS ONE v11 (2016).

31. H. Liao, et al., Phylodynamic analysis of the dissemination of HIV-1 CRF01_AE in Vietnam. Virology 391, 51–56 (2009).

32. L. N. Chhong, et al., Prevalence and clinical manifestations of dengue in older patients in Bangkok Hospital for Tropical Diseases, Thailand. Trans. R. Soc. Trop. Med. Hyg. 114, 674–681 (2020).

33. M. R. Kasper, et al., Infectious Etiologies of Acute Febrile Illness among Patients Seeking Health Care in South-Central Cambodia. Am. J. Trop. Med. Hyg. 86, 246–253 (2012).

34. K. Chheng, et al., A Prospective Study of the Causes of Febrile Illness Requiring Hospitalization in Children in Cambodia. PLOS ONE 8, e60634 (2013).

35. Cambodian Ministry of Health, National strategic plan for elimination of malaria in the Kingdom of Cambodia 2011–2025.

36. Y. Liu, et al., Evolutionary enhancement of Zika virus infectivity in Aedes aegypti mosquitoes. Nature 545, 482–486 (2017).

37. K. A. Tsetsarkin, D. L. Vanlandingham, C. E. McGee, S. Higgs, A Single Mutation in Chikungunya Virus Affects Vector Specificity and Epidemic Potential. PLOS Pathog. 3, e201 (2007).

38. F. Jourdain, et al., From importation to autochthonous transmission: Drivers of chikungunya and dengue emergence in a temperate area. PLoS Negl. Trop. Dis. 14, e0008320 (2020).

39. C. Rodriguez, et al., Fatal Encephalitis Caused by Cristoli Virus, an Emerging Orthobunyavirus, France. Emerg. Infect. Dis. 26, 1287–1290 (2020).

40. Z.-D. Wang, et al., A New Segmented Virus Associated with Human Febrile Illness in China. N. Engl. J. Med. (2019) https:/doi.org/10.1056/NEJMoa1805068 (February 2, 2021).

41. P. Pérot, et al., Identification of Umbre Orthobunyavirus as a Novel Zoonotic Virus Responsible for Lethal Encephalitis in 2 French Patients with Hypogammaglobulinemia. Clin. Infect. Dis. 72, 1701–1708 (2021).

42. N. T. T. Pham, C. T. Nguyen, H. H. Vu, Assessing and modelling vulnerability to dengue in the Mekong Delta of Vietnam by geospatial and time-series approaches. Environ. Res. 186, 109545 (2020).

43. Manning JE et al, Rapid metagenomic characterization of a case of imported COVID-19 in Cambodia. bioRvix (2020) https:/doi.org/10.1101/2020.03.02.968818 (April 17, 2020).

44. , Severe acute respiratory syndrome coronavirus 2 isolate SARS-CoV-2/human/KHM/B117/2021, complete genome (2021) (June 9, 2021).

45. M. T. Robinson, J. Satjanadumrong, T. Hughes, J. Stenos, S. D. Blacksell, Diagnosis of spotted fever group Rickettsia infections: the Asian perspective. Epidemiol. Infect. 147 (2019).

46. , PM announces plan for ‘Global Pandemic Radar.’ GOV.UK (May 24, 2021).

